# Tuberculosis prevalence among children with severe acute malnutrition: a systematic review and meta-analysis

**DOI:** 10.64898/2026.08.12.26360317

**Authors:** Aleezay A. Khan, Jasmine Armour-Marshall, Muhammad Bashir Abdullahi, Lawan Bukar, Cécile Cazes, Chishala Chabala, Mohammod Jobayer Chisti, Moussa Mamane Oumarou Farouk, Anthony J. Garcia-Prats, Catherine Hewison, Helena Huerga, Olivier Marcy, Modu Gofama Mustapha, Urhioke Ochuko, Mathew J. Reeves, Andrés Arias-Rodríguez, James A. Seddon, Tania A. Thomas, Anca Vasiliu, Bryan J. Vonasek, the Child Malnutrition and TB Working Group under The International Union Against Tuberculosis and Lung Disease

## Abstract

**Introduction:** Control of tuberculosis (TB) in children remains a major challenge globally. There is growing recognition that children with severe acute malnutrition (SAM) are a high-risk population for TB, but the global burden of TB in this group has never been comprehensively quantified.

**Methods:** We conducted a systematic review and meta-analysis to estimate the prevalence of TB among children with SAM. Following PRISMA guidelines, we searched PubMed/MEDLINE, Embase, Scopus, Web of Science, Cochrane Library, and WHO Global Index Medicus from database inception to June 15, 2026. We included studies reporting TB among systematically sampled cohorts of children <15 years with SAM as defined by the World Health Organization. Methodological study quality was assessed with adapted versions of the Newcastle-Ottawa Scale or the Joanna Briggs Institute critical appraisal checklist. Pooled TB prevalence was calculated using a random-effects model with predefined stratification of studies by geographic region, national TB incidence, and study quality. We also conducted subgroup analyses by age, sex, HIV status, SAM type, and TB exposure.

**Results:** We included 73 studies comprising 33,869 children with SAM across 15 countries, predominantly from sub-Saharan Africa and South Asia, and predominantly reporting on hospitalized children. The pooled TB prevalence was 13% (95% CI: 11-16%), but there was substantial heterogeneity (I²=98%). Studies conducted in Southern Africa had the highest pooled TB prevalence (36%, 95% CI: 19-56%) compared to other regions (p<0.01). Pooled TB prevalence was higher in those with history of TB household exposure compared to those without (74% vs. 17%, p=0.01).

**Conclusions:** Approximately one in eight children hospitalized with SAM have TB, greatest among children with history of TB exposure and those in Southern Africa. These findings highlight opportunities for improved early TB diagnosis and routine, integrated TB screening within hospital-based SAM care pathways.

## BACKGROUND

Children carry a substantial burden of tuberculosis (TB), with an estimated 1.1 million children <15 years old suffering from the disease resulting in 170,000 deaths from TB in 2024. Using World Health Organization (WHO) estimates, Southeast Asia and Africa WHO Regions have the highest prevalence of TB in the <15 years age group: about 7.3 per 100,000 (or 0.073%) [1]. Poor nutritional status can progress to secondary immunodeficiency that increases an individual’s risk of progressing to TB disease if they are infected with *M. tuberculosis*, and having severe forms of the disease and higher mortality [2–4]. There is growing recognition of the interplay between malnutrition and TB in children, and the WHO now highlights children with severe acute malnutrition (SAM) as a key high-risk group for targeted efforts for improved TB diagnosis [5,6]. SAM encompasses both severe wasting (formerly known as marasmus) and edematous malnutrition (formerly known as kwashiorkor). The WHO defines severe wasting as: weight-for-length/height z-score <-3 or mid-upper arm circumference (MUAC) <11.5 cm for those 6 to 59 months old, and Body Mass Index (BMI)-for-age z-score <-3 or MUAC-for-age z-score <-3. Edematous malnutrition is SAM characterized by the presence of bilateral pitting edema [7]. Although the global epidemiology of edematous malnutrition is not well described, in 2024 there were an estimated 12.2 million children under 5 years old with severe wasting globally, with most of these children in Asia (76%) and Africa (22%) [8].

Two recent systematic reviews have evaluated the risk for TB in the context of malnutrition. One review evaluated undernutrition (BMI <18.5 kg/m^2^ for adults) as a binary risk factor for incident TB disease. Data from all age groups were included, but most included studies only reporting on adults. Meta-analysis showed that undernutrition was associated with a relative risk of 1.96 (95% CI 1.73-2.21) for incident TB.[9] The other review evaluated the relationship between BMI as a continuous variable and TB risk exclusively in adults, using a more refined analytical approach with assessment of log-linear relationships. Meta-analysis revealed an inverse dose-response whereby decreasing BMI was associated with increasing incident TB risk, with a relative risk of 5.8 (95% CI 4.8-7.0) for BMI 16.0 kg/m^2^ versus 25 kg/m^2^ [10].

Despite the explicit biological and clinical connection between TB and SAM, the burden of TB in children with SAM is not well characterized across different settings.

Inconsistent definitions for SAM and pediatric TB diagnostic criteria, underreporting, and limited integration of TB screening into malnutrition protocols have hindered accurate estimation of the co-prevalence of these conditions [11]. Alone, TB and SAM are key drivers of preventable mortality in young children globally. When these conditions co-occur, mortality risk compounds, as demonstrated by two large studies showing that children with SAM have two to three times greater risk of mortality when they have co- morbid TB compared to those without TB [12,13]. Given the high individual and combined morbidity and mortality of TB and SAM, improved understanding of their intersection is urgently needed to inform targeted screening, diagnosis, and treatment interventions [3]. We therefore conducted a systematic review and meta-analysis to estimate the prevalence of TB among children with WHO-defined SAM receiving clinical care, with particular relevance to TB-endemic countries.

## METHODS

### Study Registration

This systematic review and meta-analysis was conducted according to the Preferred Reporting Items for Systematic Reviews and Meta-Analyses checklist [14]. The completed checklist is Table S1 in the Online Supplementary Document. The review protocol and records are available online through the Prospective Register of Systematic Reviews (PROSPERO: CRD420251057369).

### Eligibility Criteria

We included observational studies (cross-sectional and cohort) and surveillance reports that reported the prevalence of TB among children <15 years old diagnosed with SAM. Interventional studies were also included if they reported baseline prevalence of TB prior to intervention. Studies reporting on a mixed population with some not having SAM could be included if separate results were reported for only those with SAM. SAM had to be defined according to WHO criteria (see above) [7]. Children with SAM could be receiving either inpatient or outpatient care. Enumeration of children with TB could entail either robust research-based definitions or programmatic/routine diagnoses.

Studies were excluded if they did not report extractable data on TB prevalence among children with WHO-defined SAM, did not report data specifically for individuals <15 years old, or utilized study designs that did not systematically sample children with SAM.

### Data Sources and Search Strategy

A comprehensive search of the electronic databases including PubMed/MEDLINE, EMBASE, Scopus, Web of Science, Cochrane Library, and WHO Global Index Medicus was performed from database inception to June 15, 2026. Search strategy included the following terms and combinations: (‘severe acute malnutrition’ OR kwashiorkor OR ‘protein-energy malnutrition’ OR ‘nutritional edema’ OR ‘nutritional oedema’ OR marasmus OR ‘severe wasting’) AND tuberculosis. Searches using medical subject heading (MeSH) terms were conducted in PubMed, including the terms ‘severe acute malnutrition’ AND ‘tuberculosis.’ Details of the search strategy can be found in Appendix S1 in the Online Supplementary Document. To identify any relevant published or unpublished data not identified with our electronic search, 1) the reference lists of all included studies and relevant review articles were screened for potentially eligible studies, 2) we contacted experts in the field of child TB and SAM, and 3) datasets from grey literature (e.g., conference abstracts, government reports) were reviewed and considered for inclusion. No language or date restrictions were applied.

### Study Selection & Data Extraction

Two independent reviewers (AAK, JAM, AV, or BJV) screened titles and abstracts, followed by full-text review using predefined inclusion criteria. We used Covidence Systematic Review Software (Veritas Health Innovation, Melbourne, www.covidence.org) to manage the selection of studies. Data were extracted independently by two reviewers (AAK, JAM, or BJV) using a standardized form and entered into an Excel database capturing: title, first author, publication year, design type, country, summarized patient age, proportion female, number of children with SAM, number with edematous vs. non-edematous SAM, number of children with TB (including breakdown of microbiologically confirmed and clinical diagnoses without confirmation, as feasible), TB diagnostic criteria utilized, HIV prevalence, BCG immunization coverage, and history of TB exposure prevalence. Discordance between the first two reviewers on screening of titles and abstracts, review of full texts, and data on the number of children with SAM or TB was resolved by discussion amongst the reviewers or evaluation by the third reviewer.

### Assessment of Study Quality

Methodological study quality was assessed independently by two reviewers (AAK, JAM, or BJV) using the Newcastle-Ottawa Scale [15] for cohort studies and the Joanna Briggs Institute critical appraisal checklist [16] for cross-sectional studies, both tailored to this review and available in Appendix S2 the Online Supplementary Document. Prespecified score ranges for both tools were used to classify each study as ‘low,’ ‘moderate,’ or ‘high’ quality. Discrepancies in classification between the first two reviewers were resolved by discussion amongst the reviewers or evaluation by the third reviewer.

### Statistical Analysis

To estimate the pooled prevalence of TB in the included studies, we used a random effects meta-analysis with the metaprop command in Stata version 19 (StataCorp, College Station, USA). We conducted prespecified stratified meta-analyses with stratification of studies by United Nations geoscheme subregion, national TB annual incidence [1], and study quality. We planned prespecified meta-analyses of the following subgroups: HIV status, history of TB exposure, categorized age, sex, SAM type (edematous vs non-edematous), setting (e.g. inpatient vs outpatient), diagnostic method (microbiologic/confirmed vs clinical/unconfirmed), and BCG vaccination.

However, subgroup meta-analyses were only conducted if at least five included studies reported data for a particular subgroup, and therefore subgroup analyses by setting, diagnostic method, and BCG vaccination were not conducted. One study was identified as a statistical and methodologic outlier, and therefore a post-hoc leave-one-out sensitivity analyses was performed with this study excluded and meta-analyses repeated.

Prevalence estimates for each study and each pooled estimate were reported with 95% confidence intervals (CIs), with forest plots generated. Heterogeneity was assessed using the I^2^ statistic. Variation was considered to be high when I^2^ was 75% or larger [17]. Statistically significant differences were defined by p<0.05. Because heterogeneity was very high, we did not evaluate for publication bias [18].

## RESULTS

### Study Selection

Figure 1 depicts the study selection process. A total of 504 records were identified through database searches. Title and abstract screening included 380 records, 167 full-text articles were assessed for eligibility, and 69 were retained after full-text review. An additional four studies were identified outside the search so that 73 studies were included in total.

**Figure 1.**
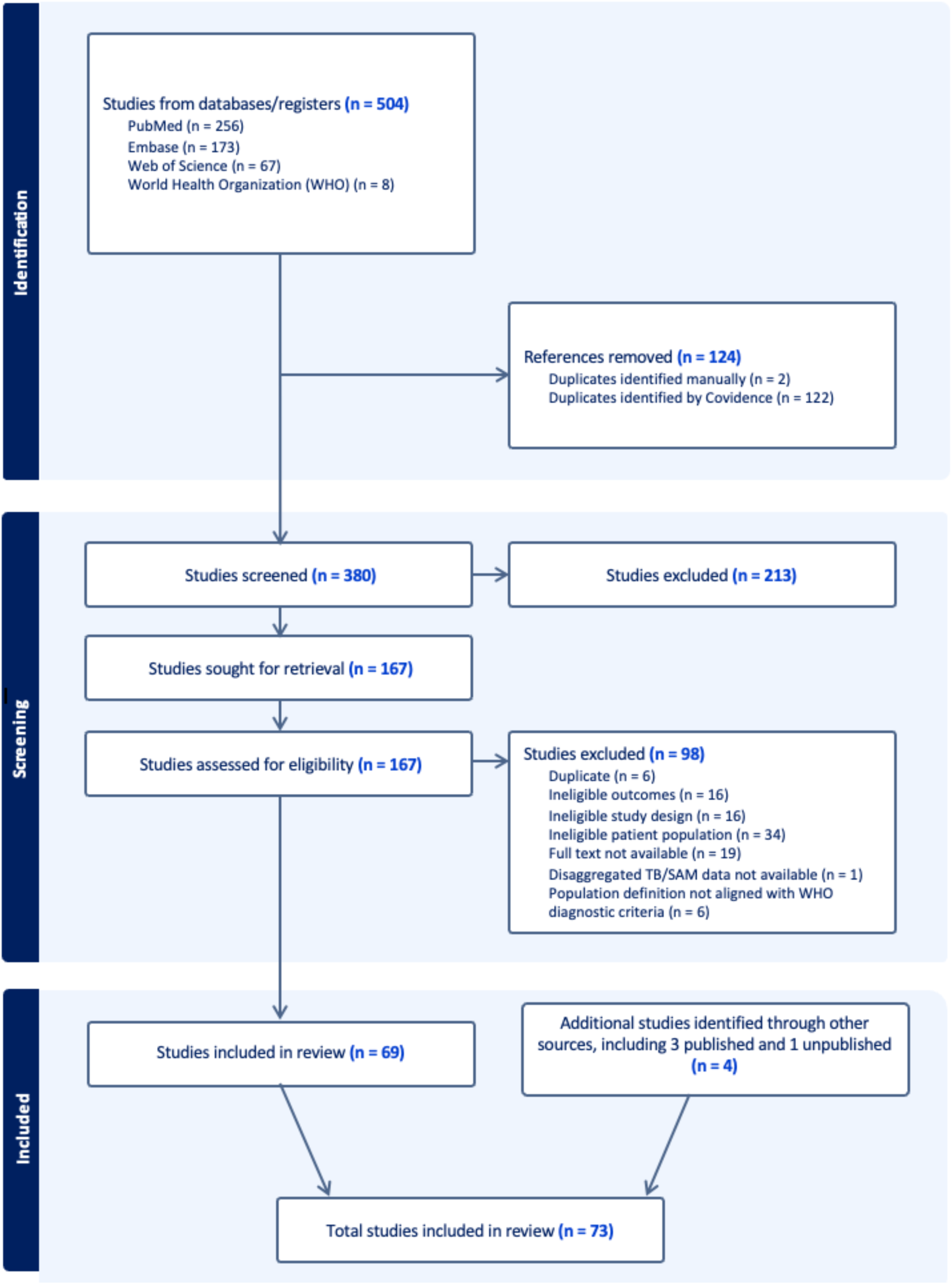
PRISMA flow diagram of study selection.

### Study Characteristics

The 73 included studies reported on a total of 33,869 children with SAM, with study sizes ranging from 45 to 9540 participants. All studies were conducted in sub-Saharan Africa or South Asia. One study was reported in French (Clavier-Rogez 2015), and all other studies were reported in English. All the studies were observational in design, including 32 cohort studies and 41 cross-sectional studies. Studies were conducted between 2003 and 2025, with most studies conducted in the past decade. All studies were conducted primarily in inpatient hospital-based settings, though one cohort study reported on an extensive follow-up period mostly in the outpatient setting after TB diagnosis was made during hospitalization (Clavier-Rogez 2015) and the TB ALGO PED study recruited 3% of the cohort from outpatient facilities and 97% from inpatient facilities. Most participants were less than five years old, with reported median or mean ages ranging from 10 to 36 months. Among studies reporting sex distribution, the proportion of female participants ranged from 16% to 66%, with most studies demonstrating a relatively balanced distribution. Studies reported HIV prevalences ranging from 0% to 51%. For study quality assessments, 17 (23%) studies were classified as high quality, 28 (38%) as moderate quality, and 28 (38%) as low quality.

There were 8 (11%) studies conducted in TB high-incidence countries (national annual incidence >250 per 100,000), 34 (47%) studies in medium-incidence countries (150 to 250 per 100,000), and 31 (42%) studies in low-incidence countries (<150 per 100,000). Study characteristics are summarized in Table 1.

**Table 1:**
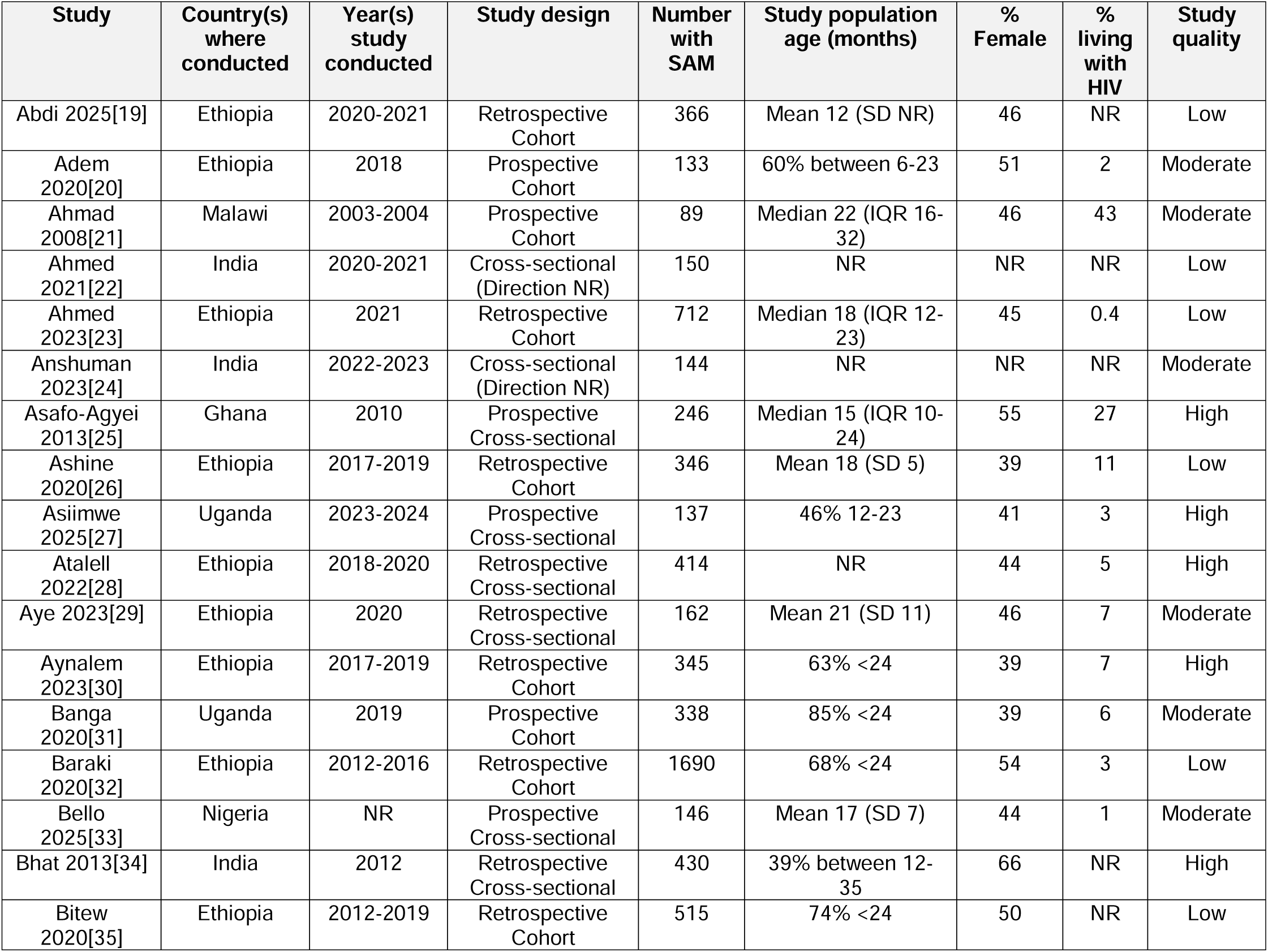
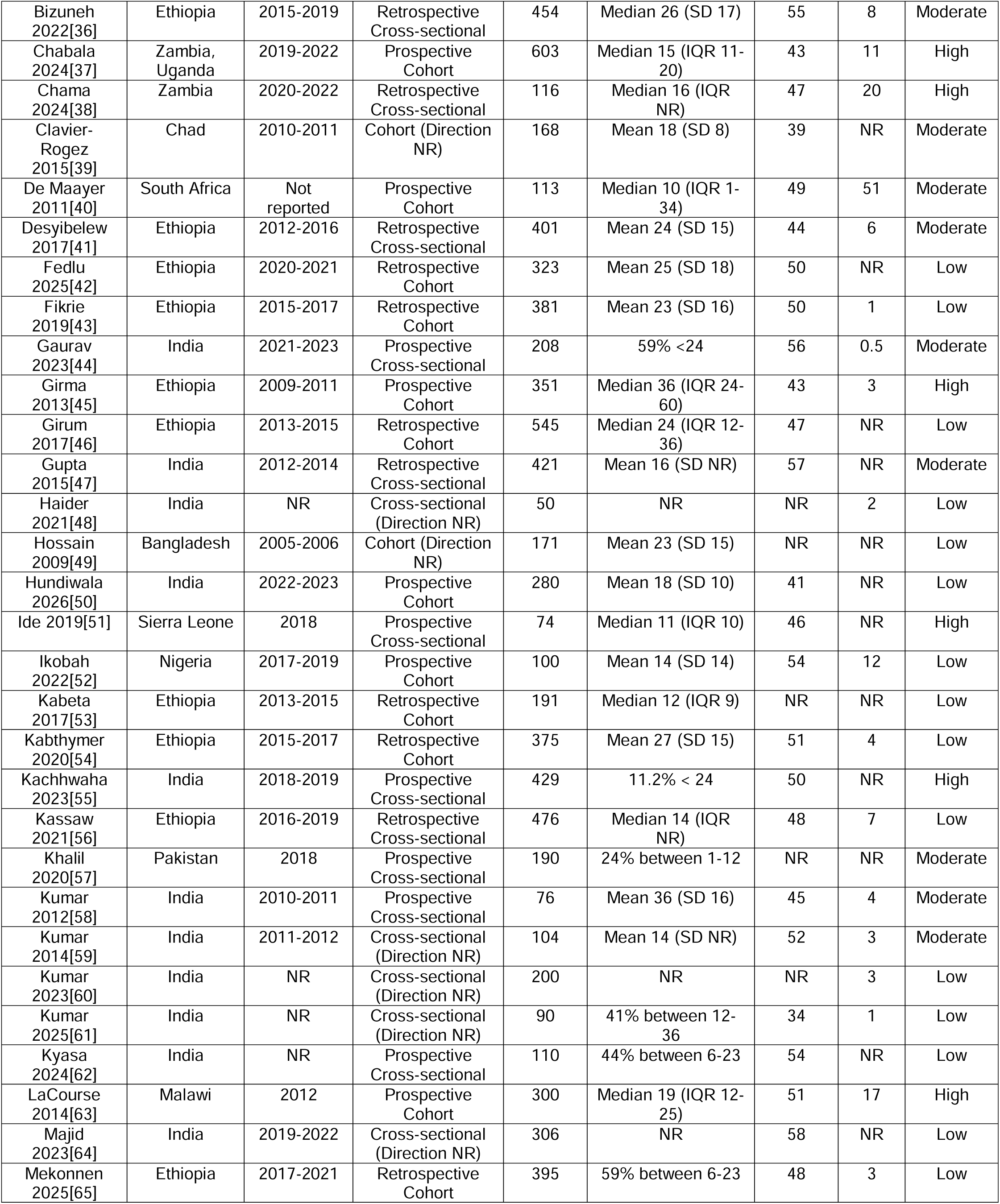
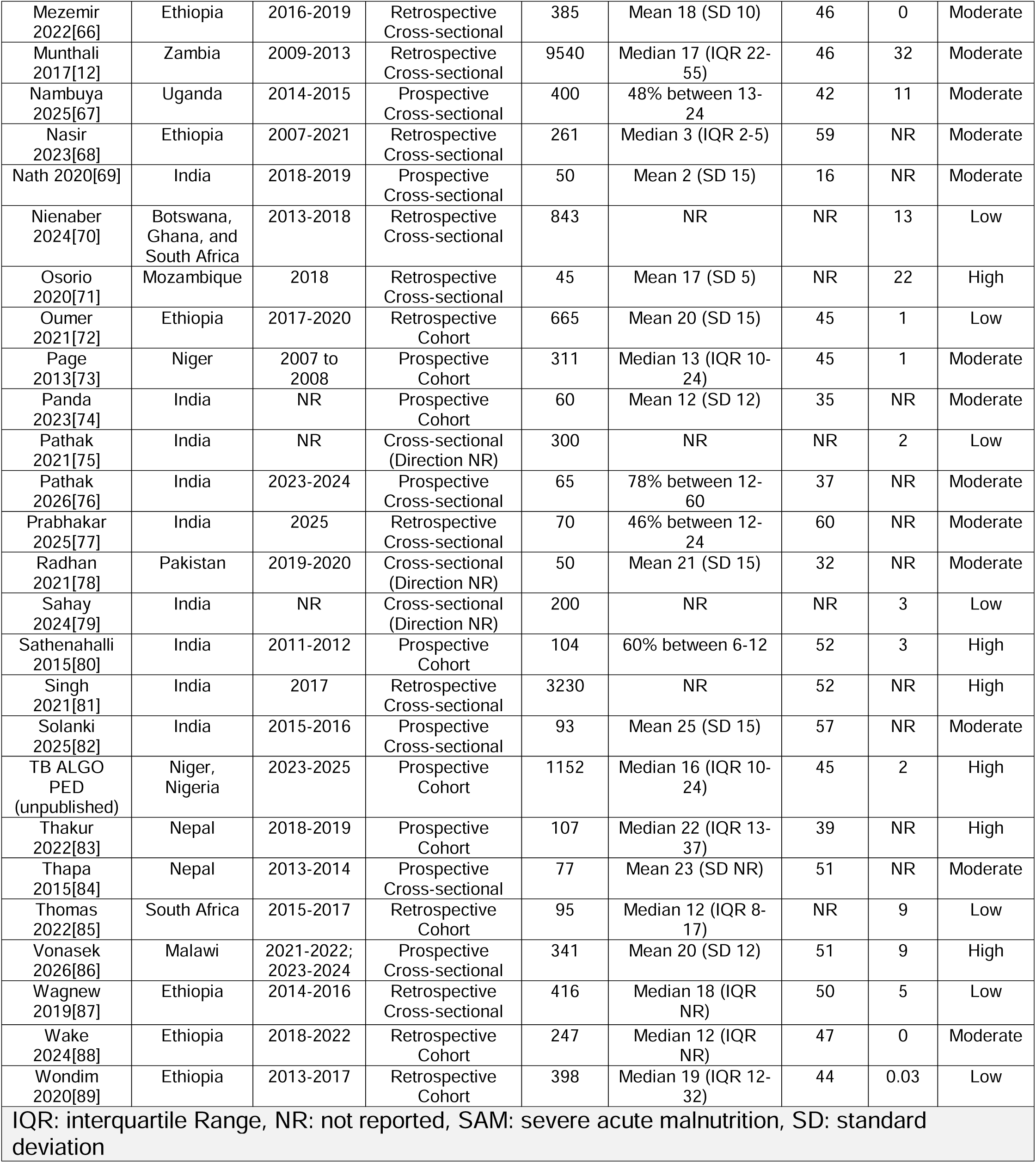
Characteristics of the included studies.

### Meta-analyses

The pooled prevalence of TB among children with SAM from all included studies was 13% (95% CI 11-16%) but with high heterogeneity (I^2^ = 98%, p<0.01) and study-specific prevalences ranging from 1% (95% CI 1-3%) to 73% (95% CI 68-78%), as shown in Figure 2.

**Figure 2.**
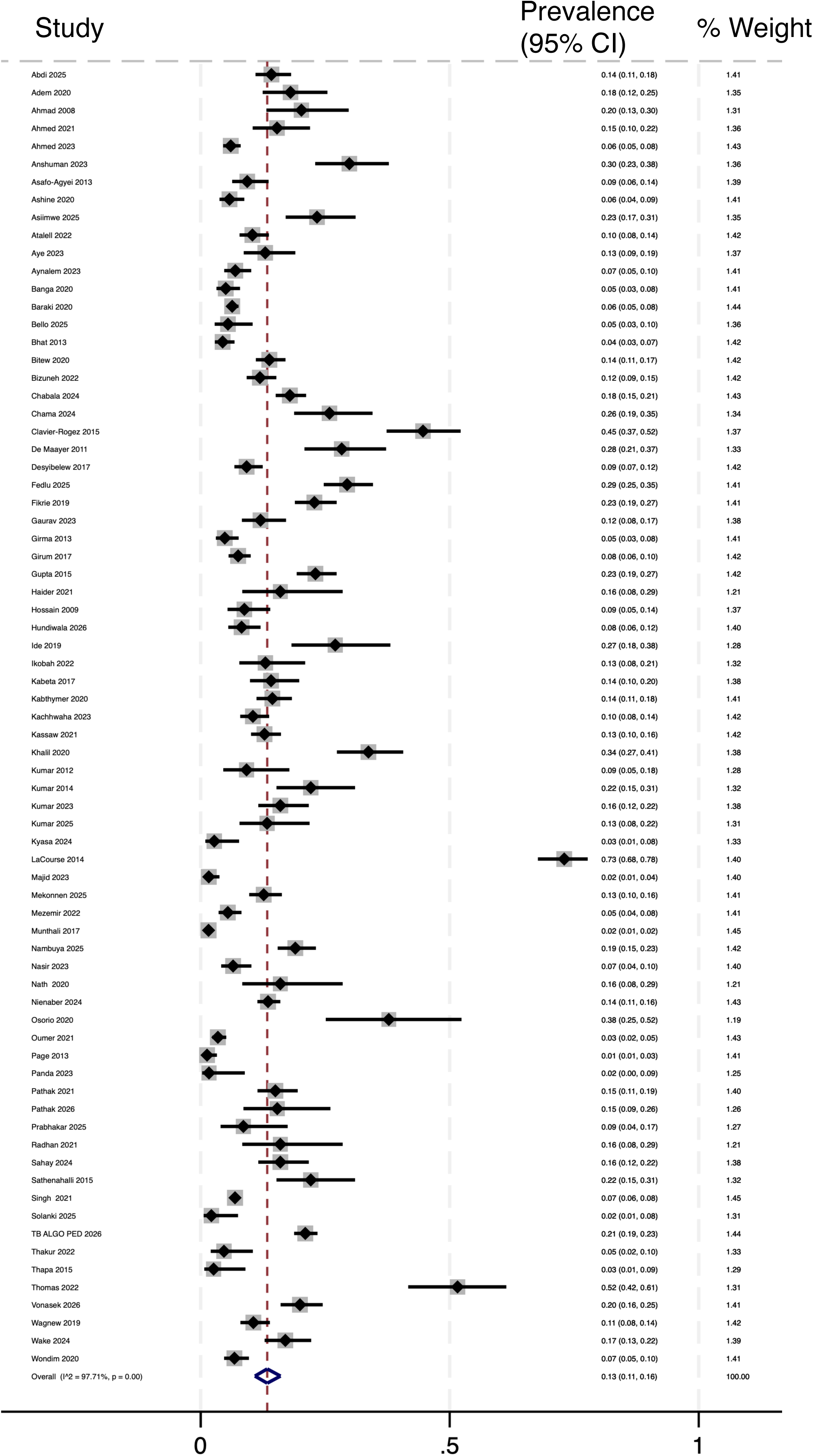
Forest plot of meta-analysis of the prevalence of tuberculosis in children with severe acute malnutrition for all included studies. CI: confidence interval.

Meta-analyses with studies stratified by United Nations geoscheme subregion, study quality, and national TB annual incidence are summarized in Table 2 and detailed in forest plots in Figures S1, S2, and S3 in the Online Supplementary Document, respectively. The pooled TB prevalence was significantly higher (p<0.001) for studies conducted in Southern Africa (36%, 95% CI 19-56%) compared to other subregions.

**Table 2.** Pooled prevalence of TB among children with SAM by stratified and subgroup meta-analyses.

|  | **N** | **Prevalence (95% CI)** | **$I^2$** | ***p*-value** |
| --- | --- | --- | --- | --- |
| All studies | 73 | 13% (11-16%) | 98% | - |
| <u>UN geoscheme subregion</u> |  |  |  |  |
| Eastern Africa | 28 | 11% (9-13%) | 93% | <0.001 |
| Southern Africa | 7 | 36% (19-56%) | 98% |  |
| Southern Asia | 29 | 11% (8-15%) | 97% |  |
| Western Africa | 7 | 15% (6-27%) | 97% |  |
| <u>Study quality</u> |  |  |  |  |
| Low | 28 | 12% (9-15%) | 94% | 0.331 |
| Moderate | 28 | 13% (8-18%) | 98% |  |
| High | 17 | 17% (11-24%) | 98% |  |
| <u>Nation annual TB incidence</u> |  |  |  |  |
| Low (<150 per 100,000) | 31 | 13% (10-17%) | 97% | 0.285 |
| Medium (150-250 per 100,000) | 34 | 11% (9-14%) | 93% |  |
| High (>250 per 100,000) | 8 | 26% (8-48%) | 99% |  |
| <u>HIV status</u> |  |  |  |  |
| Infected | 7 | 43% (11-78%) | 99% | 0.080 |
| Negative | 6 | 12% (3-26%) | 99% |  |
| <u>History of TB exposure</u> |  |  |  |  |
| None | 6 | 17% (5-35%) | 99% | 0.009 |
| Present | 6 | 74% (33-99%) | 97% |  |
| <u>Age</u> |  |  |  |  |
| <24 months | 8 | 11% (7-15%) | 83% | 0.297 |
| ≥24 months | 8 | 15% (8-22%) | 81% |  |
| <u>Sex</u> |  |  |  |  |
| Female | 12 | 20% (8-36%) | 99% | 0.797 |
| Male | 12 | 18% (7-32%) | 99% |  |
| <u>SAM type</u> |  |  |  |  |
| No nutritional edema | 7 | 24% (13-38%) | 97% | 0.986 |
| Nutritional edema present | 7 | 24% (10-42%) | 96% |  |
| CI: confidence interval, N: number of studies in the meta-analysis, SAM: severe acute malnutrition, TB: tuberculosis, UN: United Nations |  |  |  |  |

There was no significant variation in pooled TB prevalence after stratifying by study quality (p=0.331) or national TB annual incidence (p=0.285). Heterogeneity was high across all strata with I^2^ of 93% or higher (all with p<0.01).

Meta-analyses for subgroups defined by participant HIV status, history of TB exposure, age, sex, and SAM type are summarized in Table 2 and detailed in forest plots in Figures S4, S5, S6, S7, and S8 in the Online Supplementary Document, respectively. There was a trend towards higher pooled TB prevalence for those living with HIV, but the difference was not statistically significant (43% vs 12%, p=0.08).

Pooled TB prevalence was higher for those with history of TB exposure (74% vs 17%, p=0.009). Heterogeneity was high across all subgroups with I^2^ of 80% or higher (all with p<0.01).

One study (LaCourse 2014) was identified as a statistical and methodologic outlier (detailed in the Discussion below). Repeated meta-analyses with this study excluded are shown in Table S2 in the Online Supplementary Document. The overall pooled prevalence was still 13% with a slightly narrower 95% CI (11-15%), and there were no notable changes to results from the meta-analyses with stratification of studies or subgroups.

## DISCUSSION

In this systematic review of TB prevalence in children with WHO-defined SAM the pooled prevalence was strikingly high at 13% (95% CI: 11–16%). This review included data from over thirty thousand children, almost all of whom were hospitalized with SAM. All included studies were conducted in sub-Saharan Africa or South Asia, which reflects the regions where most children with SAM reside [8] and where over half of the global burden of TB is estimated to occur [1]. TB prevalence was highly heterogeneous across studies. Subgroup analyses did not demonstrate key drivers of the heterogeneity, but were notable for significantly higher TB prevalence in those with a history of TB exposure and a trend towards higher prevalence in those living with HIV, though that trend was not statistically significant. To our knowledge, this is the first systematic review of TB prevalence in children with SAM.

Malnutrition and TB share a bidirectional biological relationship, resulting in the high prevalence of TB in children with SAM that we report in this review. Malnutrition impairs both innate and adaptive immunity, increasing susceptibility to TB [2,90]. In the other direction of this relationship, TB also exacerbates malnutrition [90]. Compounding the biologic connections between malnutrition and TB are social and economic factors that promote both conditions. TB and SAM tend to occur most frequently in marginalized populations of children living in poverty with poor access to health care services.

Our findings may underestimate the true burden of TB in children hospitalized with SAM. Most of the data in this review are from children less than five years old, the age group most affected by SAM. According to the WHO, an estimated 58% of children in this age group with TB globally were not diagnosed or reported to have TB in 2022 [5]. Particularly for the included retrospective studies (N=32), reliance on routine diagnosis of TB may have led to underestimation of TB prevalence. A major component of our study quality assessments in this review was the robustness of the TB diagnostic criteria, and 38% of included studies were ‘low’ quality largely because of limited description of the TB in the manuscript or reliance on routine diagnosis and reporting of TB. With more intensive diagnostic workup under research conditions and using more rigorous TB diagnostic criteria, fewer missed diagnoses and higher estimation of TB prevalence in young children could be expected. However, when we stratified pooled TB prevalence estimates by the three study quality categories, no differences were observed.

The wide range of individual study estimates of TB prevalence (1–73%), which translated into high heterogeneity (I² = 98%), was expected given the diversity of settings studied, differences in study design, and different TB diagnostic approaches. TB epidemiology varies widely between and within countries [1]. Surprisingly, when we stratified pooled TB prevalence estimates by categories of national TB incidence for the countries in which individual studies were conducted, no significant differences were observed. On the other hand, we did find differences in pooled TB prevalence when stratifying studies by the region in which they were conducted, with the highest pooled prevalence (36%) for studies conducted in Southern Africa. The study by LaCourse and colleagues had a prevalence estimate of 73% [63], which was 21% more than the point prevalence of the next highest study. This study was the only included study to utilize the first version of the NIH clinical diagnosis definition for child TB [91], while several included studies utilized the revised case definitions published in 2015 [92]. A key change to the case definitions was a shift from five categories (“confirmed,” “probable,” “possible,” “unlikely,” and “not” TB) to three (“confirmed,” “unconfirmed,” and “unlikely”). In the study by LaCourse and colleagues, 66% of participants were classified as “possible” TB [63]. In the binary classification of TB status used in this review, we classified those with “possible” TB as children with TB. Had we only defined children with TB as those classified as “confirmed” and “probable” TB, the TB prevalence from that study would have been much lower at 7%. This scenario exemplifies both the challenge of defining children with TB, even with well-designed clinical studies, and the impact differences in study designs and diagnostic definitions had on the heterogeneous prevalence estimates we report here. Sensitivity analyses with this study excluded did not result in notable changes to the overall pooled TB prevalence estimate or the meta-analyses with stratification of studies or subgroups.

Active screening strategies for TB in children have traditionally focused on two high-risk groups in low- and middle-income countries: close contacts of individuals diagnosed with pulmonary TB and children living with HIV. This review highlights another important TB high-risk group, children hospitalized with SAM, which aligns with the WHO’s new emphasis in recent guidelines for implementing targeted efforts for improved TB diagnosis alongside malnutrition care [5,6]. Children with SAM and HIV— doubling up on risk factors—had an especially high pooled TB prevalence of 43%. In the sub-group analysis, this prevalence was not significantly higher compared to HIV-negative children, and the lack of difference was driven by a large single study that reported very low TB prevalence in both children living with HIV and HIV-negative children (2% and 1%, respectively) [12]. Especially in sub-Saharan Africa, pediatric HIV infection remains unacceptably high, and our data highlight the deadly synergy between HIV, SAM, and TB.

Children with SAM and history of TB exposure—also doubling up on risk factors—had a strikingly high TB prevalence of 74% in subgroup analysis pooling data from six studies. We note the impact of incorporation bias with this association, as history of TB exposure influences the clinical diagnosis of TB in children. Nonetheless, clinicians caring for children with SAM should always inquire about TB exposure, and when this history is present, there should be a low threshold for initiating treatment for TB disease. Using the recently developed WHO-recommended treatment decision algorithms for child TB, there is a clear recommendation to treat for TB disease when a child with SAM has a history of TB exposure [6], and our data support this approach.

The care pathways for management of SAM are longitudinal and offer serial opportunities to assess for TB or establish alternative diagnoses. This programmatic advantage can be leveraged for improved screening and diagnosis of TB, especially if there is strong coordination between TB and malnutrition programs (which otherwise tend to be siloed) to provide integrated services that can benefit children with co-prevalent TB and SAM the most.

The notable strengths of this systematic review and meta-analysis are the large numbers of studies and children included and the restriction to only include studies reporting on children with WHO-defined SAM, which allowed for a consistently defined population that is programmatically relevant. There are noteworthy limitations with this study. First, as detailed above, the TB diagnostic criteria varied amongst the included studies, and the high proportion of ‘low’ quality included studies was driven by weaker diagnostic criteria. Second, 66% of the studies were from only two countries, Ethiopia and India. Settings with high burden of SAM and TB globally may not be well represented. Third, data on outpatients with SAM were essentially absent. Over the past few decades, guidelines and programs have shifted so that children with SAM receive outpatient, community-based care when they lack medical complications or danger signs [7], and therefore most children with SAM nowadays are initially managed as outpatients. We therefore highlight a need for primary research on the burden of TB in children receiving SAM care as outpatients.

## CONCLUSIONS

In summary, our results demonstrate the high burden of TB in children hospitalized with SAM. These results emphasize the urgent need for improved strategies to diagnose TB tailored to this unique population. Building upon the treatment decision algorithms for child TB newly recommended by the WHO in 2022 [6], SAM-specific algorithms could optimize accuracy [37]. Quantifying the high prevalence of TB in children hospitalized with SAM informs policymakers and public health officials about a key population to target for TB case finding efforts and emphasizes to clinicians the importance of having a high index of suspicion for TB when caring for children with SAM.

## Data Availability

All study materials (template data collection forms, data extracted from included studies, data used for analyses, analytic code, and any other materials) are available from the corresponding author upon reasonable request.

## Acknowledgements

We thank the authors of the primary studies included in this review.

## Author contributions

JAM, LB, CC, CC, MJC, MMOF, AJGP, CH, HH, OM, MGM, UO, JAS, TAT, AV, and BJV conceptualized the study. AAK drafted and finalized the study protocol with input from JAM, LB, CC, CC, MJC, MMOF, AJGP, CH, HH, OM, MGM, UO, JAS, TAT, AV, and BJV. AAK, JAM, AV, and BJV assessed articles for inclusion, extracted data, and performed methodologic quality assessments. AAK and BJV analyzed the data, and drafted and finalized the manuscript. MJR and AAR gave essential guidance on data analysis. All authors gave input on interpretation of the analyses, critically reviewed the manuscript, and approved the final version.

## Availability of Study Materials

All study materials (template data collection forms, data extracted from included studies, data used for analyses, analytic code, and any other materials) are available from the corresponding author upon request.

## Supplementary Figures

**Figure S1.** Forest plot of meta-analysis of the prevalence of tuberculosis in children with severe acute malnutrition with studies stratified by United Nations geoscheme subregion. CI: confidence interval.

**Figure S2.** Forest plot of meta-analysis of the prevalence of tuberculosis in children with severe acute malnutrition with studies stratified by quality. CI: confidence interval.

**Figure S3.** Forest plot of meta-analysis of the prevalence of tuberculosis in children with severe acute malnutrition with studies stratified by national tuberculosis annual incidence (low: <150 per 100,000; medium: 150 to 250 per 100,000; high: >250 per 100,000). CI: confidence interval.

**Figure S4.** Forest plot of meta-analysis of the prevalence of tuberculosis in children with severe acute malnutrition—sub-group analysis by HIV status. CI: confidence interval.

**Figure S5.** Forest plot of meta-analysis of the prevalence of tuberculosis in children with severe acute malnutrition—sub-group analysis by presence or absence of history of exposure to an individual with tuberculosis. CI: confidence interval.

**Figure S6.** Forest plot of meta-analysis of the prevalence of tuberculosis in children with severe acute malnutrition—sub-group analysis by age. CI: confidence interval.

**Figure S7.** Forest plot of meta-analysis of the prevalence of tuberculosis in children with severe acute malnutrition—sub-group analysis by sex. CI: confidence interval.

**Figure S8.** Forest plot of meta-analysis of the prevalence of tuberculosis in children with severe acute malnutrition—sub-group analysis by presence or absence of nutritional edema. CI: confidence interval.

