## Supplementary Document for "Tuberculosis prevalence among children with severe acute malnutrition: a systematic review and meta-analysis"

### Table S1. PRISMA 2020 Checklist

| Section & Topic | Item # | Checklist item | Location where item is reported |
|---|---|---|---|
| **TITLE** | | | |
| Title | 1 | Identify the report as a systematic review. | p. 1 |
| **ABSTRACT** | | | |
| Abstract | 2 | See the PRISMA 2020 for Abstracts checklist. | p. 3 |
| **INTRODUCTION** | | | |
| Rationale | 3 | Describe the rationale for the review in the context of existing knowledge. | p. 4 & 5 |
| Objectives | 4 | Provide an explicit statement of the objective(s) or question(s) the review addresses. | p. 5 |
| **METHODS** | | | |
| Eligibility criteria | 5 | Specify the inclusion and exclusion criteria for the review and how studies were grouped for the syntheses. | p. 5 & 6 |
| Information sources | 6 | Specify all databases, registers, websites, organisations, reference lists and other sources searched or consulted to identify studies. Specify the date when each source was last searched or consulted. | p. 5 |
| Search strategy | 7 | Present the full search strategies for all databases, registers and websites, including any filters and limits used. | Table S2 |
| Selection process | 8 | Specify the methods used to decide whether a study met the inclusion criteria of the review, including how many reviewers screened each record and each report retrieved, whether they worked independently, and if applicable, details of automation tools used in the process. | p. 6 |
| Data collection process | 9 | Specify the methods used to collect data from reports, including how many reviewers collected data from each report, whether they worked independently, any processes for obtaining or confirming data from study investigators, and if applicable, details of automation tools used in the process. | p. 6 |
| Data items | 10a | List and define all outcomes for which data were sought. Specify whether all results that were compatible with each outcome domain in each study were sought (e.g. for all measures, time points, analyses), and if not, the methods used to decide which results to collect. | p. 6 |
| | 10b | List and define all other variables for which data were sought (e.g. participant and intervention characteristics, funding sources). Describe any assumptions made about any missing or unclear information. | p. 6 |
| Study risk of bias assessment | 11 | Specify the methods used to assess risk of bias in the included studies, including details of the tool(s) used, how many reviewers assessed each study and whether they worked independently, and if applicable, details of automation tools used in the process. | p. 6 |
| Effect measures | 12 | Specify for each outcome the effect measure(s) (e.g. risk ratio, mean difference) used in the synthesis or presentation of results. | p. 6 |
| Synthesis methods | 13a | Describe the processes used to decide which studies were eligible for each synthesis (e.g. tabulating the study intervention characteristics and comparing against the planned groups for each synthesis (item #5)). | p. 6 |
| | 13b | Describe any methods required to prepare the data for presentation or synthesis, such as handling of missing summary statistics, or data conversions. | p. 6 |
| | 13c | Describe any methods used to tabulate or visually display results of individual studies and syntheses. | p. 6 |
| | 13d | Describe any methods used to synthesize results and provide a rationale for the choice(s). If meta-analysis was performed, describe the model(s), method(s) to identify the presence and extent of statistical heterogeneity, and software package(s) used. | p. 6 |
| | 13e | Describe any methods used to explore possible causes of heterogeneity among study results (e.g. subgroup analysis, meta-regression). | p. 6 |
| | 13f | Describe any sensitivity analyses conducted to assess robustness of the synthesized results. | p. 6 |
| Reporting bias assessment | 14 | Describe any methods used to assess risk of bias due to missing results in a synthesis (arising from reporting biases). | p. 6 |
| Certainty assessment | 15 | Describe any methods used to assess certainty (or confidence) in the body of evidence for an outcome. | p. 6 |

#### Table S1. PRISMA 2020 Checklist

| Section & Topic | Item # | Checklist item | Location where item is reported |
|---|---|---|---|
| **RESULTS** | | | |
| Study selection | 16a | Describe the results of the search and selection process, from the number of records identified in the search to the number of studies included in the review, ideally using a flow diagram. | Figure 1 |
| | 16b | Cite studies that might appear to meet the inclusion criteria, but which were excluded, and explain why they were excluded. | Figure 1 |
| Study characteristics | 17 | Cite each included study and present its characteristics. | Table 1 |
| Risk of bias in studies | 18 | Present assessments of risk of bias for each included study. | Table 1 |
| Results of individual studies | 19 | For all outcomes, present, for each study: (a) summary statistics for each group (where appropriate) and (b) an effect estimate and its precision (e.g. confidence/credible interval), ideally using structured tables or plots. | Figure 2 |
| Results of syntheses | 20a | For each synthesis, briefly summarise the characteristics and risk of bias among contributing studies. | p. 7 |
| | 20b | Present results of all statistical syntheses conducted. If meta-analysis was done, present for each the summary estimate and its precision (e.g. confidence/credible interval) and measures of statistical heterogeneity. If comparing groups, describe the direction of the effect. | p. 10 & 11 |
| | 20c | Present results of all investigations of possible causes of heterogeneity among study results. | p. 10 & 11 |
| | 20d | Present results of all sensitivity analyses conducted to assess the robustness of the synthesized results. | p. 10 & 11 |
| Reporting biases | 21 | Present assessments of risk of bias due to missing results (arising from reporting biases) for each synthesis assessed. | n/a |
| Certainty of evidence | 22 | Present assessments of certainty (or confidence) in the body of evidence for each outcome assessed. | p. 10 |
| **DISCUSSION** | | | |
| Discussion | 23a | Provide a general interpretation of the results in the context of other evidence. | p. 11 & 12 |
| | 23b | Discuss any limitations of the evidence included in the review. | p. 13 & 14 |
| | 23c | Discuss any limitations of the review processes used. | p. 13 & 14 |
| | 23d | Discuss implications of the results for practice, policy, and future research. | p. 13 & 14 |
| **OTHER INFORMATION** | | | |
| Registration and protocol | 24a | Provide registration information for the review, including register name and registration number, or state that the review was not registered. | p. 5 |
| | 24b | Indicate where the review protocol can be accessed, or state that a protocol was not prepared. | p. 5 |
| | 24c | Describe and explain any amendments to information provided at registration or in the protocol. | n/a |
| Support | 25 | Describe sources of financial or non-financial support for the review, and the role of the funders or sponsors in the review. | p. 14 |
| Competing interests | 26 | Declare any competing interests of review authors. | p. 14 |
| Availability of data, code and other materials | 27 | Report which of the following are publicly available and where they can be found: template data collection forms; data extracted from included studies; data used for all analyses; analytic code; any other materials used in the review. | p. 14 |

*From:* Page MJ, McKenzie JE, Bossuyt PM, Boutron I, Hoffmann TC, Mulrow CD, et al. The PRISMA 2020 statement: an updated guideline for reporting systematic reviews. BMJ 2021;372:n71. doi: 10.1136/bmj.n71. This work is licensed under CC BY 4.0. To view a copy of this license, visit https://creativecommons.org/licenses/by/4.0/

### Appendix S1. Search Strategy from Selected Databases

#### Scopus

Advanced query

( ( tuberculosis MeSH Terms ) AND ( "severe acute malnutrition" MeSH Terms ) ) OR ( ( ( "severe acute malnutrition" Text Word ) OR ( kwashiorkor Text Word ) OR ( "protein-energy malnutrition" Text Word ) OR ( marasmus Text Word ) OR ( "severe wasting" Text Word ) OR ( "nutritional oedema" Text Word ) OR ( "nutritional edema" Text Word ) ) AND ( tuberculosis Text Word ) )

Show less

Edit in advanced search

Documents | Preprints (Beta) | Secondary documents

Are you searching for: ( ( tuberculosis[MeSH Terms] ) AND ( "severe acute malnutrition"[MeSH Terms] ) ) OR ( ( ...

21 documents found

#### PubMed

##### History and Search Details

Download | Delete

| Search | Actions | Details | Query | Results | Time |
|---|---|---|---|---|---|
| #1 | ••• | ! > | Search: **((tuberculosis[MeSH Terms]) AND ("severe acute malnutrition"[MeSH Terms])) OR (((((("severe acute malnutrition"[Text Word]) OR (kwashiorkor[Text Word])) OR ("protein-energy malnutrition"[Text Word])) OR (marasmus[Text Word])) OR ("severe wasting"[Text Word])) OR ("nutritional oedema"[Text Word])) OR ("nutritional edema"[Text Word])) AND (tuberculosis[Text Word]))** Sort by: **Most Recent** | 257 | 07:43:35 |

#### Web of Science

**102** results from Web of Science Core Collection for:

(TS=(severe acute malnutrition)) AND TS=(tuberculosis)

Copy query link

+ Add Keywords    Quick add keywords: + severe acute malnutrition | + survival status | + severe malnutrition | + retrospective cohort | + wast

**102** Documents    You may also like...    Analyze Results | Citation Report | Create Alert

#### Embase

Enter search query e.g. 'cancer gene therapy'/exp OR ((treatment OR therapy) NEAR/5 fluorouracil):ab

('tuberculosis'/exp AND 'severe acute malnutrition'/exp OR (('severe acute malnutrition' OR kwashiorkor OR 'protein-energy malnutri

Mapping | Date | Fields | Quick limits

Search history

Select all | Combine using AND | Save | Delete | Export | Email

| | | | |
|---|---|---|---|
| #1 | ('tuberculosis'/exp AND 'severe acute malnutrition'/exp OR (('severe acute malnutrition' OR kwashiorkor OR 'protein-energy malnutrition' OR marasmus OR 'severe wasting') AND tuberculosis)) AND 'severe acute malnutrition' AND 'tuberculosis' | 172 | ⋮ |

172 results for search #1    Create email alert    Index miner

#### APPENDIX S2. RISK-OF-BIAS AND STUDY-QUALITY ASSESSMENTS

##### Cohort studies

The adapted Newcastle-Ottawa Scale contained six assessment items. Points were additive, and criteria that were not met or not reported received 0 points. Possible total scores ranged from 0 to 9.

**Assessment criteria and scoring**

| No. | Assessment item | Scoring rule | Max. |
|---|---|---|---|
| 1 | **Representativeness of the population at risk** | **1 point:** Children with SAM were recruited from a representative setting (multicenter, community, outpatient, or inpatient hospital).<br>**1 point:** Systematic sampling was clearly described (e.g., consecutive sampling or interval sampling of every X children). | 2 |
| 2 | **Ascertainment of SAM status** | **1 point:** The SAM definition was clearly stated in the article.<br>**0 points:** SAM was not clearly defined. | 1 |
| 3 | **Outcome presence (TB diagnosis timing)** | **1 point:** TB was diagnosed at the time of SAM diagnosis or prospectively from that point, with no pre-existing TB.<br>**0 points:** Timing was not clearly described or pre-existing TB was possible. | 1 |
| 4 | **Study comparability: factors available for subgroup analyses** | **0 points:** No subgroup factors were available.<br>**1 point:** One factor was available.<br>**2 points:** Two or more factors were available.<br>**Examples:** age, HIV status, TB contact, BCG status, sex, setting, and season. | 2 |
| 5 | **Assessment of the TB outcome** | **1 point:** Any detail was provided about how participants were assessed for TB.<br>**1 point:** TB was confirmed using microbiology and/or standardized clinical criteria (e.g., Graham et al., 2015). | 2 |
| 6 | **Follow-up duration** | **1 point:** Follow-up lasted 2 months or longer to confirm the TB diagnosis.<br>**0 points:** Follow-up lasted less than 2 months or was unclear. | 1 |

**Interpretation of the total score (maximum = 9)**

| Total score | Methodological quality | Corresponding risk of bias |
|---|---|---|
| **7-9** | High | Low |
| **4-6** | Moderate | Moderate |
| **0-3** | Low | High |

**Cross-sectional studies**

The adapted Joanna Briggs Institute assessment contained eight items. For every item, Yes = 1 and No = 0, yielding possible total scores from 0 to 8.

**Assessment criteria and scoring**

| No. | Assessment item | Max. |
|---|---|---|
| **1** | Were the criteria for inclusion in the sample clearly defined? | **1** |
| **2** | Were the study participants and setting described in detail? | **1** |
| **3** | Was SAM clearly defined? | **1** |
| **4** | Was it clear that children with SAM were systematically sampled (e.g., consecutive sampling or interval sampling of every X children)? | **1** |
| **5** | Was any detail provided about how study participants were assessed for TB? | **1** |
| **6** | Were TB case definitions based on microbiologic and/or standardized clinical criteria (e.g., Graham et al.)? | **1** |
| **7** | Was at least one factor available for subgroup analyses (e.g., age, HIV status, TB contact, BCG status, sex, setting, or season)? | **1** |
| **8** | Was more than one factor available for subgroup analyses (e.g., age, HIV status, TB contact, BCG status, sex, setting, or season)? | **1** |

**Interpretation of the total score (maximum = 8)**

| Total score | Methodological quality | Corresponding risk of bias |
|---|---|---|
| **6-8** | High | Low |
| **3-5** | Moderate | Moderate |
| **0-2** | Low | High |

**Abbreviations.** BCG, Bacillus Calmette-Guerin;; SAM, severe acute malnutrition; TB, tuberculosis.

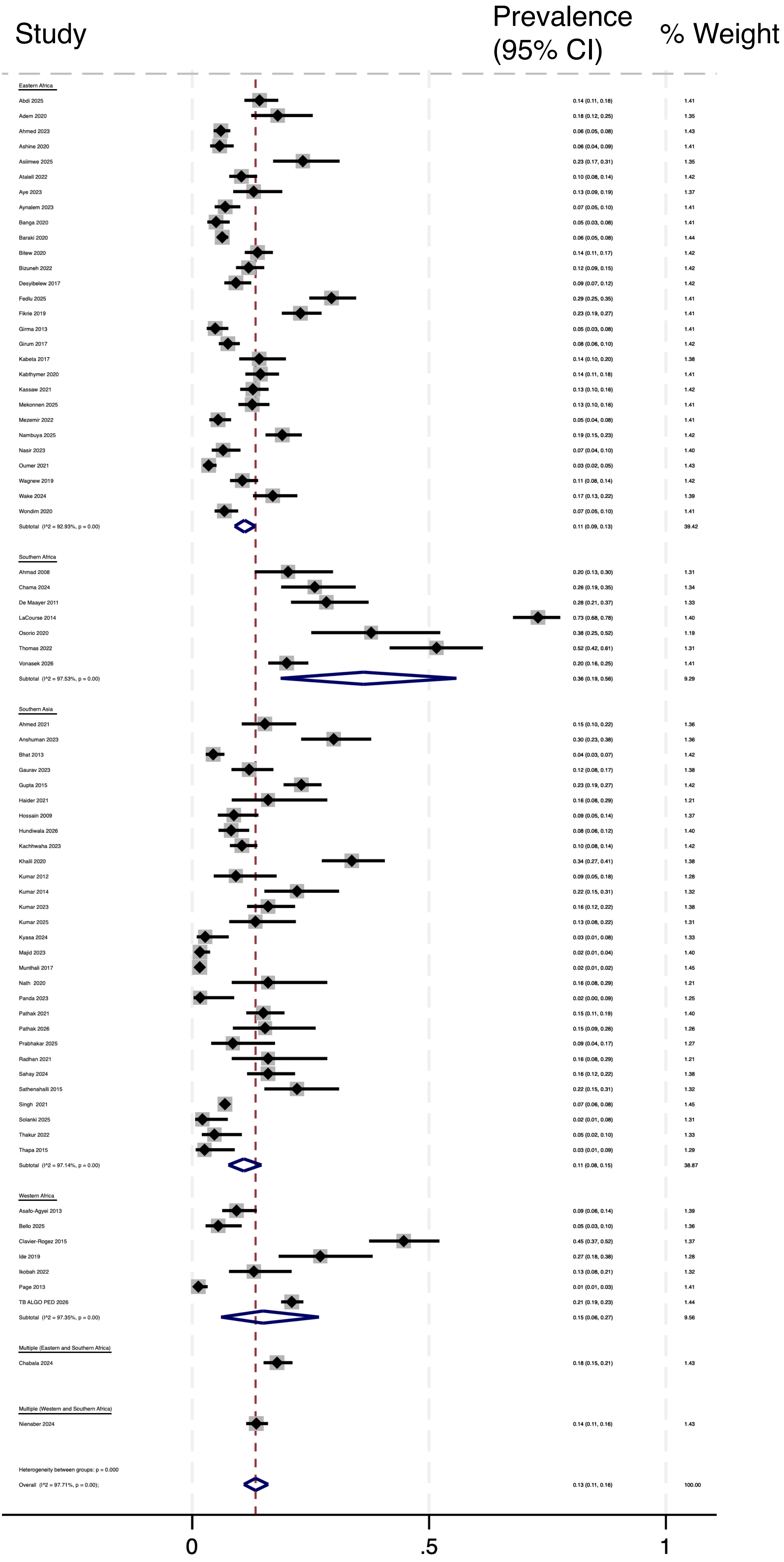


**Figure S1.** Forest plot of meta-analysis of the prevalence of tuberculosis in children with severe acute malnutrition with studies stratified by United Nations geoscheme subregion. CI: confidence interval.

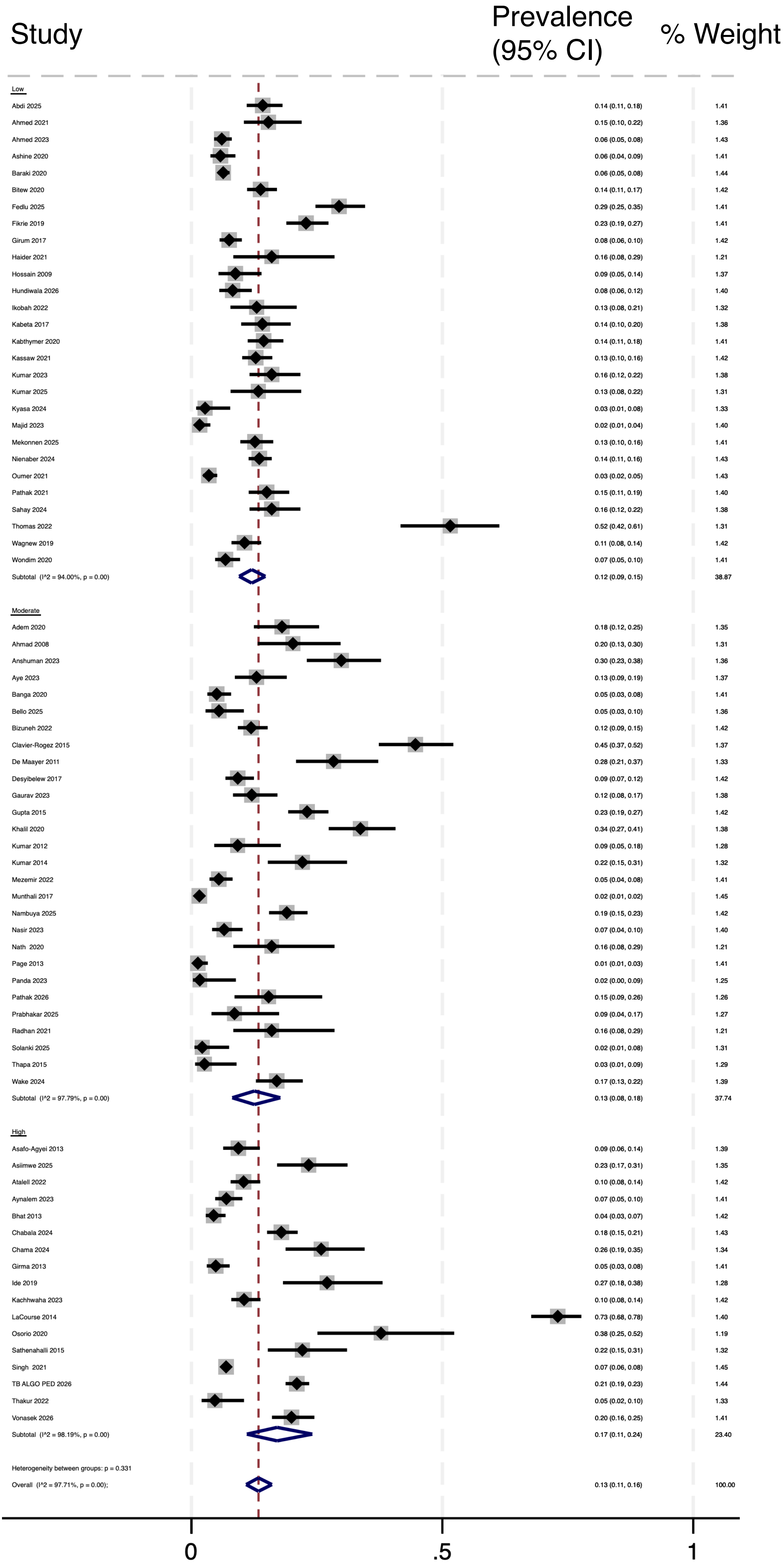


**Figure S2.** Forest plot of meta-analysis of the prevalence of tuberculosis in children with severe acute malnutrition with studies stratified by quality. CI: confidence interval.

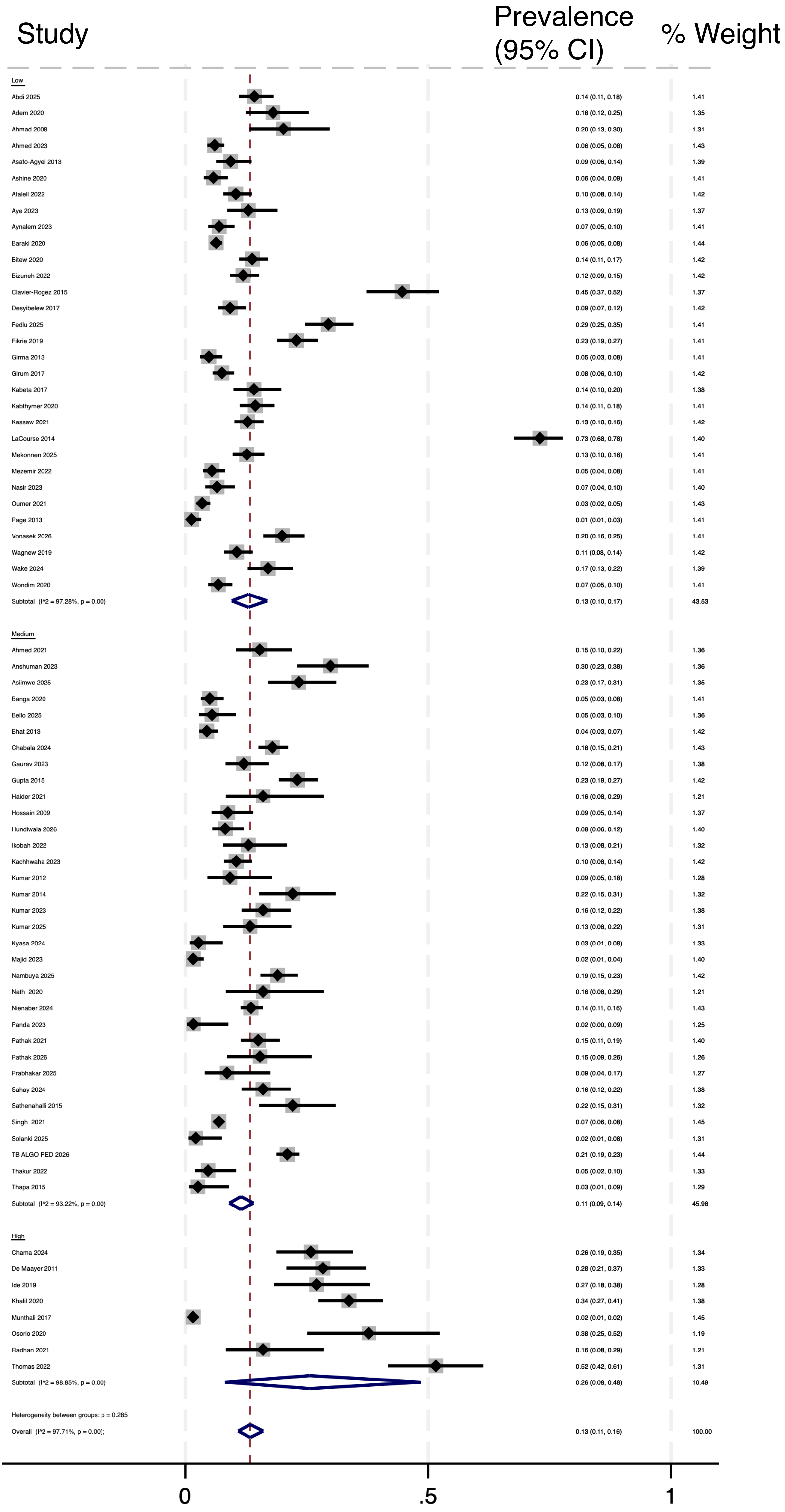


**Figure S3.** Forest plot of meta-analysis of the prevalence of tuberculosis in children with severe acute malnutrition with studies stratified by national tuberculosis annual incidence (low: <150 per 100,000; medium: 150 to 250 per 100,000; high: >250 per 100,000). CI: confidence interval.

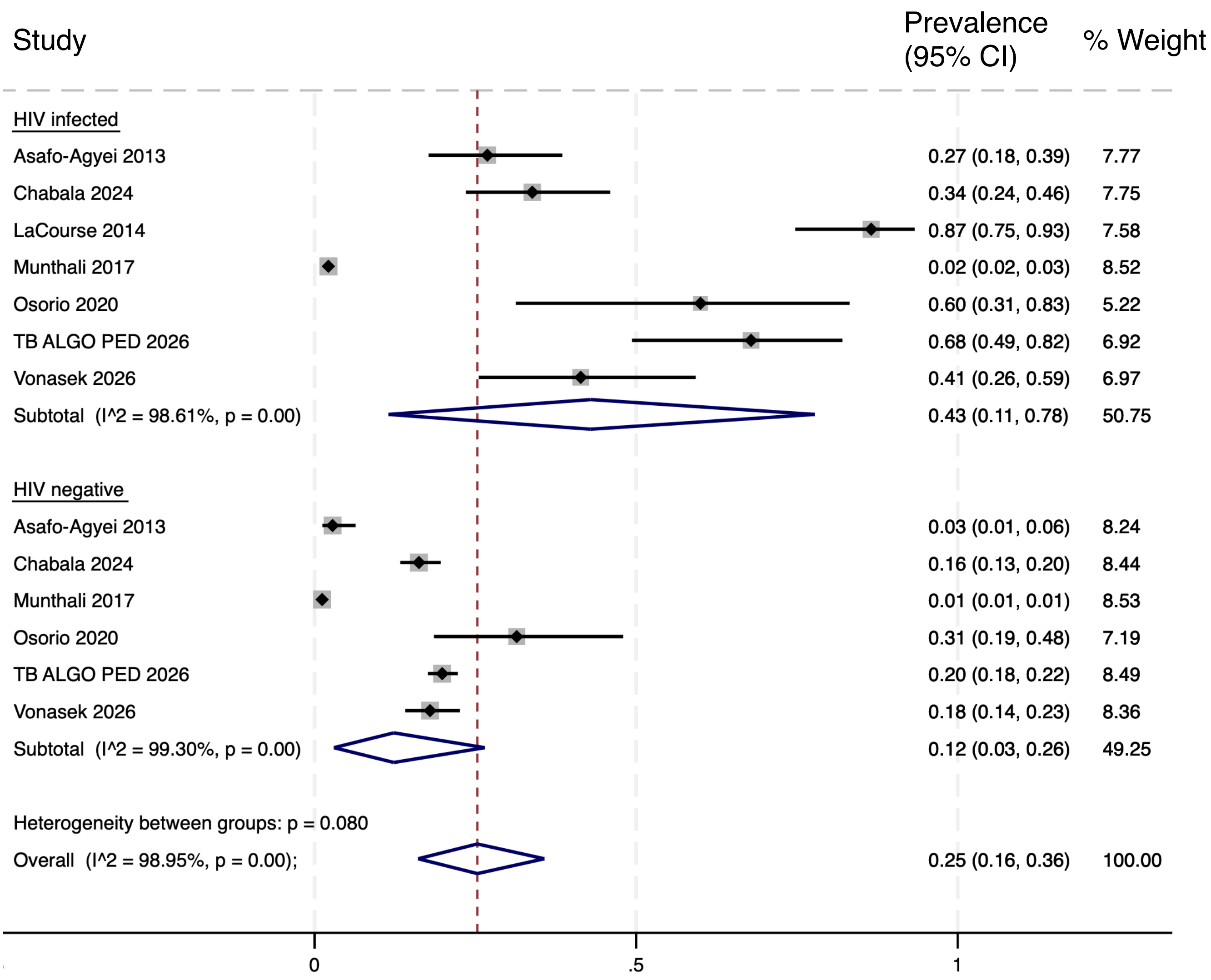


**Figure S4.** Forest plot of meta-analysis of the prevalence of tuberculosis in children with severe acute malnutrition—sub-group analysis by HIV status. CI: confidence interval.

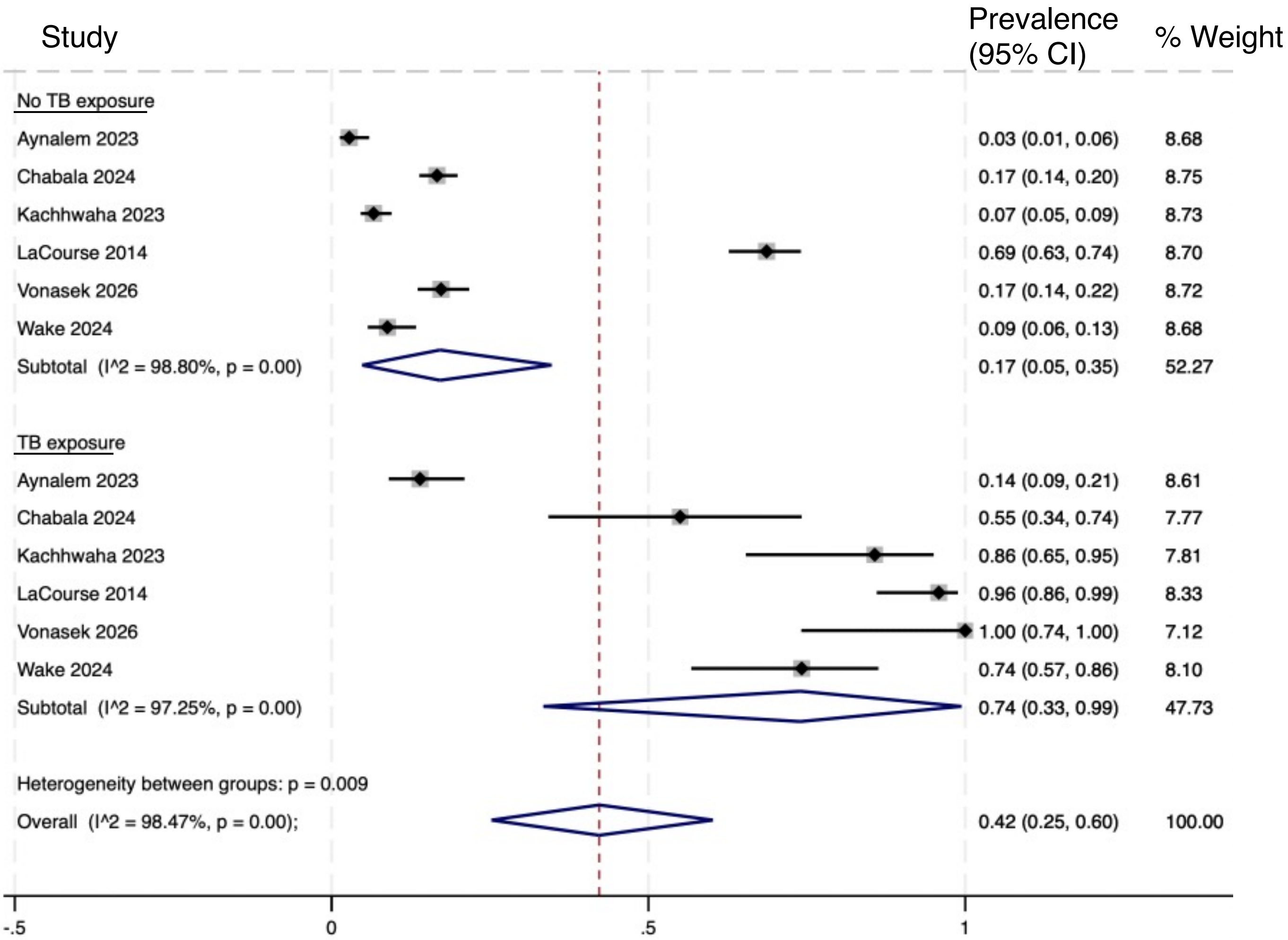


**Figure S5.** Forest plot of meta-analysis of the prevalence of tuberculosis in children with severe acute malnutrition—sub-group analysis by presence or absence of history of exposure to an individual with tuberculosis. CI: confidence interval.

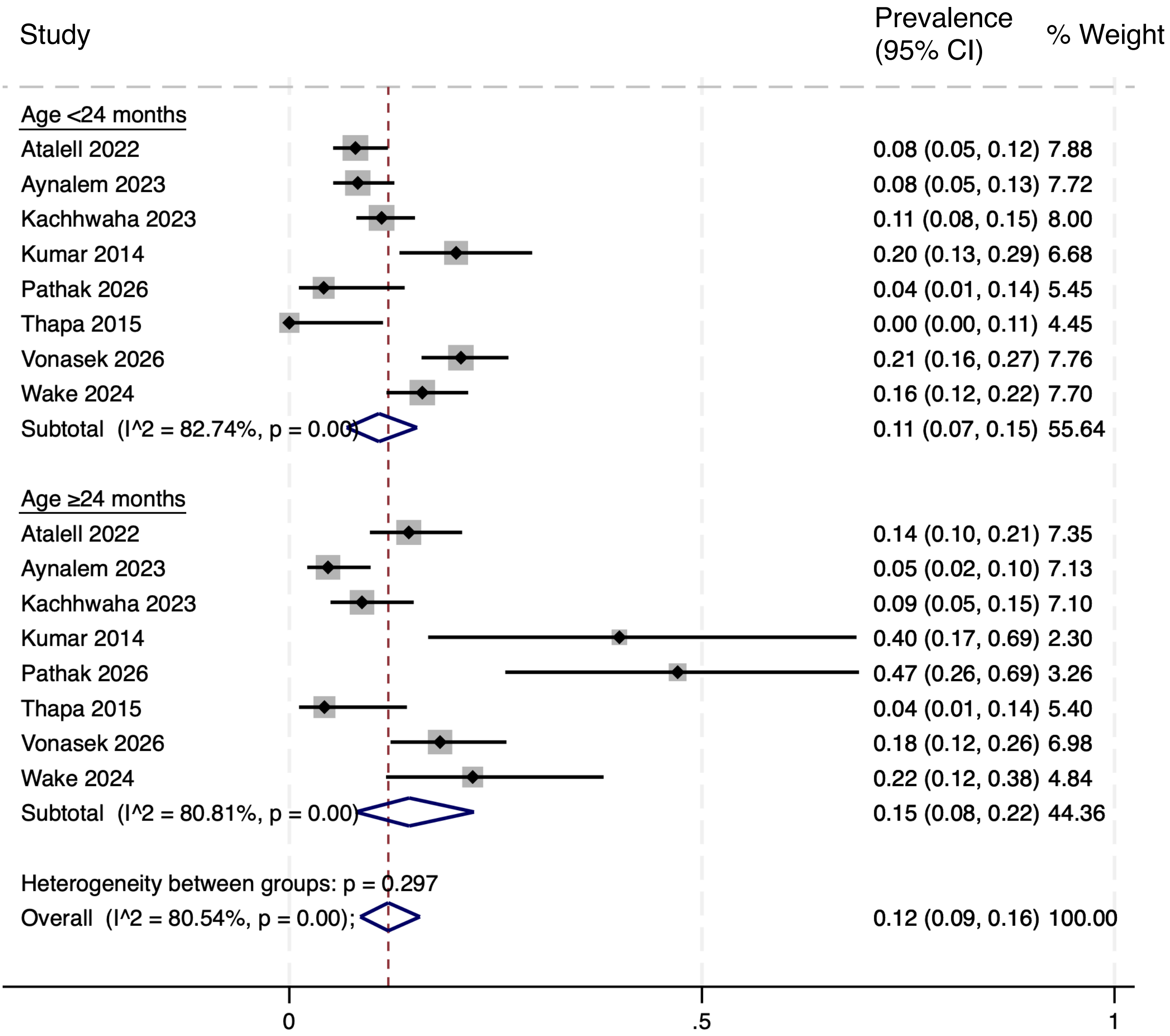


**Figure S6.** Forest plot of meta-analysis of the prevalence of tuberculosis in children with severe acute malnutrition—sub-group analysis by age. CI: confidence interval.

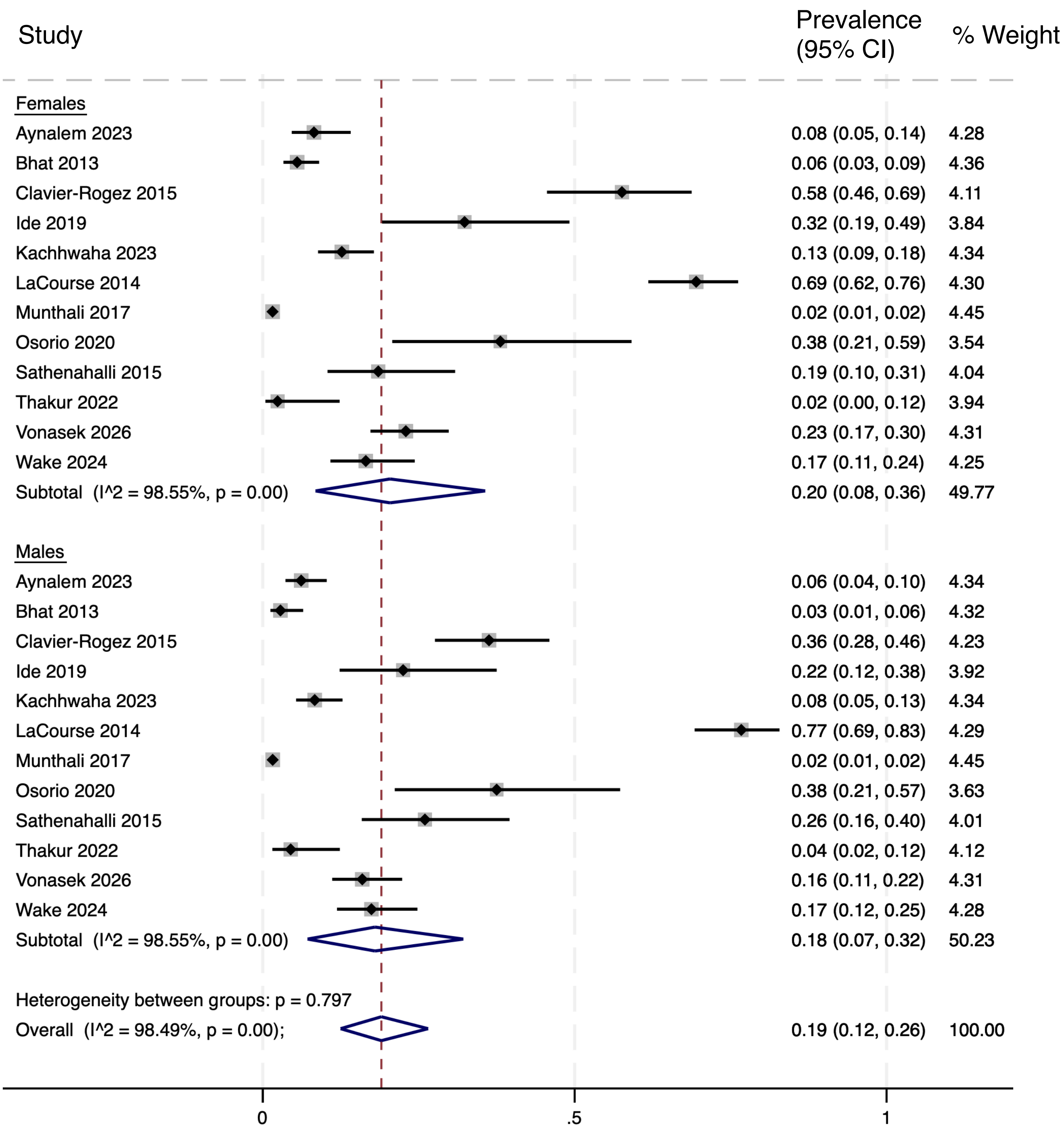


**Figure S7.** Forest plot of meta-analysis of the prevalence of tuberculosis in children with severe acute malnutrition—sub-group analysis by sex. CI: confidence interval.

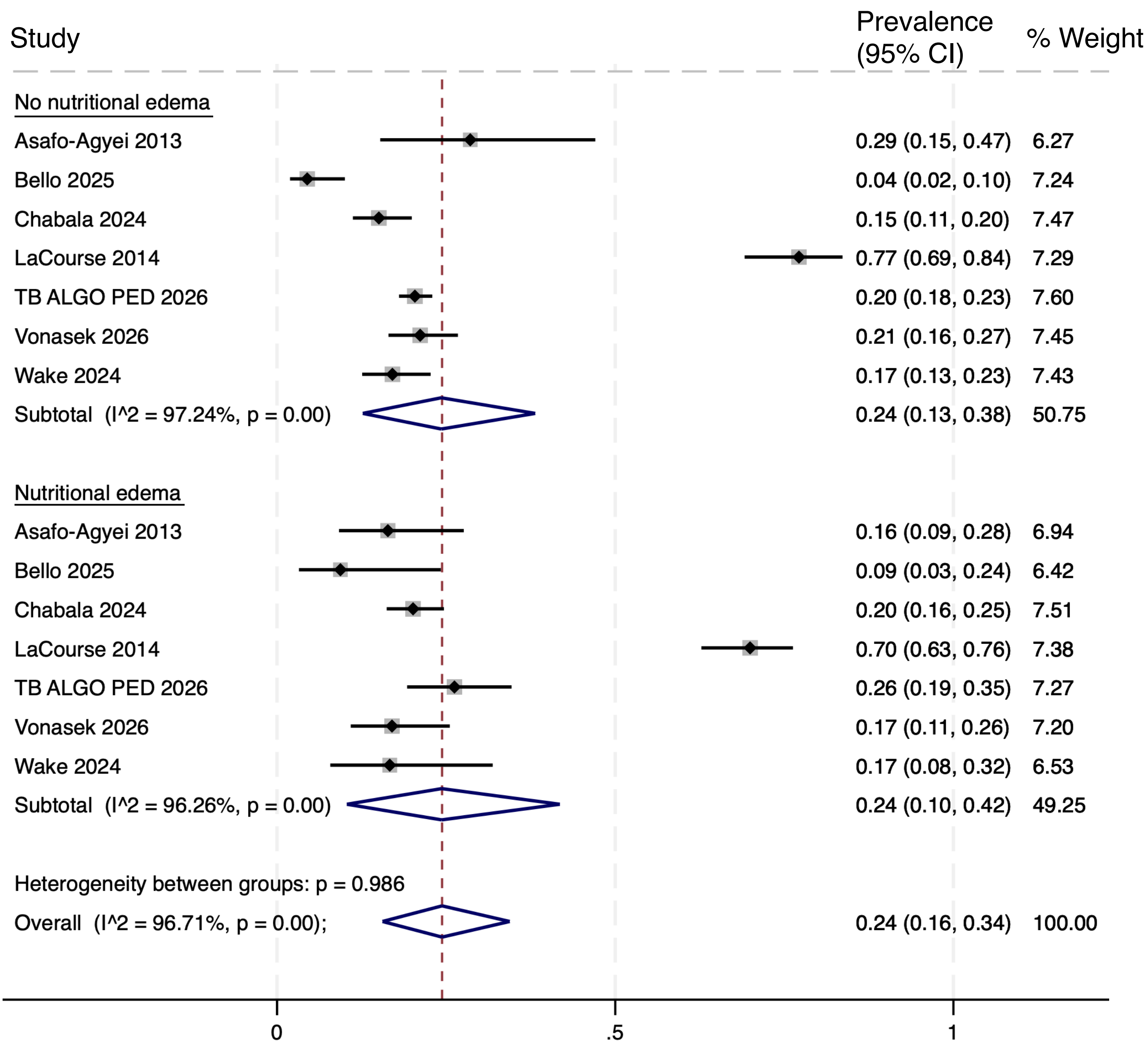


**Figure S8.** Forest plot of meta-analysis of the prevalence of tuberculosis in children with severe acute malnutrition—sub-group analysis by presence or absence of nutritional edema. CI: confidence interval.

**Table S2.** Sensitivity analysis of pooled prevalence of TB among children with SAM by stratified and subgroup meta-analyses with a single study outlier (LaCourse 2014) included and excluded.

| | **Outlier Study Included** *(same as Table 2)* | | | | **Outlier Study Excluded** | | | |
|---|---|---|---|---|---|---|---|---|
| | **N** | **Prevalence (95% CI)** | **$I^2$** | ***p*-value** | **N** | **Prevalence (95% CI)** | **$I^2$** | ***p*-value** |
| All studies | 73 | 13% (11-16%) | 98% | - | 72 | 13% (11-15%) | 97% | - |
| UN geoscheme subregion | | | | | | | | |
| Eastern Africa | 28 | 11% (9-13%) | 93% | <0.001 | 28 | 11% (9-13%) | 93% | <0.001 |
| Southern Africa | 7 | 36% (19-56%) | 98% | | 6 | 30% (21-40%) | 87% | |
| Southern Asia | 29 | 11% (8-15%) | 97% | | 29 | 11% (8-15%) | 97% | |
| Western Africa | 7 | 15% (6-27%) | 97% | | 7 | 15% (6-27%) | 97% | |
| Study quality | | | | | | | | |
| Low | 28 | 12% (9-15%) | 94% | 0.331 | 28 | 12% (9-15%) | 94% | 0.702 |
| Moderate | 28 | 13% (8-18%) | 98% | | 28 | 13% (8-18%) | 98% | |
| High | 17 | 17% (11-24%) | 98% | | 16 | 14% (10-18%) | 96% | |
| Nation annual TB incidence | | | | | | | | |
| Low (<150 per 100,000) | 31 | 13% (10-17%) | 97% | 0.285 | 30 | 11% (9-14%) | 94% | 0.331 |
| Medium (150-250 per 100,000) | 34 | 11% (9-14%) | 93% | | 34 | 11% (9-14%) | 93% | |
| High (>250 per 100,000) | 8 | 26% (8-48%) | 99% | | 8 | 26% (8-48%) | 99% | |
| HIV status | | | | | | | | |
| Infected | 7 | 43% (11-78%) | 99% | 0.080 | 6 | 35% (9-66%) | 98% | 0.136 |
| Negative | 6 | 12% (3-26%) | 99% | | 6 | 12% (3-26%) | 99% | |
| History of TB exposure | | | | | | | | |
| None | 6 | 17% (5-35%) | 99% | 0.009 | 5 | 10% (5-16%) | 93% | 0.003 |
| Present | 6 | 74% (33-99%) | 97% | | 5 | 68% (27-98%) | 96% | |
| Age | | | | | | | | |
| <24 months | 8 | 11% (7-15%) | 83% | 0.297 | - | - | - | - |
| ≥24 months | 8 | 15% (8-22%) | 81% | | | | | |
| Sex | | | | | | | | |
| Female | 12 | 20% (8-36%) | 99% | 0.797 | 11 | 16% (8-27%) | 97% | 0.633 |
| Male | 12 | 18% (7-32%) | 99% | | 11 | 13% (6-22%) | 97% | |
| SAM type | | | | | | | | |
| No nutritional edema | 7 | 24% (13-38%) | 97% | 0.986 | 6 | 16% (12-22%) | 83% | 0.349 |
| Nutritional edema present | 7 | 24% (10-42%) | 96% | | 6 | 19% (16-23%) | 19% | |

CI: confidence interval, N: number of studies in the meta-analysis, SAM: severe acute malnutrition, TB: tuberculosis, UN: United Nations
